# Genomic and epigenomic analysis of plasma cell-free DNA identifies stemness features associated with worse survival in *AR*-altered lethal prostate cancer

**DOI:** 10.1101/2023.12.01.23299215

**Authors:** Pradeep S. Chauhan, Irfan Alahi, Savar Sinha, Alexander L. Shiang, Ryan Mueller, Jace Webster, Ha X. Dang, Debanjan Saha, Lilli Greiner, Breanna Yang, Gabris Ni, Elisa M. Ledet, Ramandeep K. Babbra, Wenjia Feng, Peter K. Harris, Faridi Qaium, Ellen B. Jaeger, Patrick J. Miller, Sydney A. Caputo, Oliver Sartor, Russell K. Pachynski, Christopher A. Maher, Aadel A. Chaudhuri

**Affiliations:** Division of Cancer Biology, Department of Radiation Oncology, Washington University School of Medicine, St. Louis, Missouri, United States of America; Department of Computer Science and Engineering, Washington University in St. Louis, St. Louis, Missouri, United States of America; Division of Urology, Department of Surgery, Washington University School of Medicine, St. Louis, Missouri, United States of America; Department of Internal Medicine, Washington University School of Medicine, St. Louis, Missouri, United States of America; McDonnell Genome Institute, Washington University in St. Louis, Missouri, United States of America; Siteman Cancer Center, Washington University in St. Louis, Missouri, United States of America; Tulane University School of Medicine, New Orleans, Louisiana, United States of America; Wilmot Institute Cancer Center, University of Rochester Medical Center, Rochester, New York, United States of America; Department of Medicine, Mayo Clinic, Rochester, Minnesota, United States of America; Department of Urology, Mayo Clinic, Rochester, Minnesota, United States of America; Department of Radiology, Mayo Clinic, Rochester, Minnesota, United States of America; Mayo Clinic Comprehensive Cancer Center, Rochester, Minnesota, United States of America; Department of Biomedical Engineering, Washington University in St. Louis, St. Louis, Missouri, United States of America; Department of Genetics, Washington University School of Medicine, St. Louis, Missouri, United States of America

**Author notes:** Corresponding Author: Aadel A. Chaudhuri, M.D., Ph.D. Division of Cancer Biology, Department of Radiation Oncology Washington University School of Medicine, 4511 Forest Park Avenue, St. Louis, MO 63108. These authors contributed equally to this work (Pradeep S. Chauhan, Irfan Alahi). These authors jointly supervised this work (Oliver Sartor, Russell K. Pachynski, Christopher A. Maher, Aadel A. Chaudhuri).

**Keywords:** lethal prostate cancer, mCRPC, plasma, cell-free DNA, epigenomics, fragmentomics, methylation, stemness, next-generation sequencing, androgen receptor

## Abstract

Metastatic castration-resistant prostate cancer (mCRPC) resistant to androgen receptor (AR)-targeted agents is often lethal. Unfortunately, biomarkers for this deadly disease remain under investigation, and underpinning mechanisms are ill-understood. Here, we applied deep sequencing to ∼100 mCRPC patients prior to the initiation of first-line AR-targeted therapy, which detected *AR*/enhancer alterations in over a third of patients, which correlated with lethality. To delve into the mechanism underlying why these patients with cell-free *AR*/enhancer alterations developed more lethal prostate cancer, we next performed genome-wide cell-free DNA epigenomics. Strikingly, we found that binding sites for transcription factors associated with developmental stemness were nucleosomally more accessible. These results were corroborated using cell-free DNA methylation data, as well as tumor RNA sequencing from a held-out cohort of mCRPC patients. Thus, we validated the importance of *AR*/enhancer alterations as a prognostic biomarker in lethal mCRPC, and showed that the underlying mechanism for lethality involves reprogramming developmental states toward increased stemness.

## Introduction

AR-directed therapies such as abiraterone and enzalutamide have significantly improved survival for men with metastatic castration-resistant prostate cancer (mCRPC).^1-4^ However, approximately one-third of patients develop lethal prostate cancer that does not respond well to treatment, with very little known about determinants of resistance in this more lethal form of mCRPC.^5,6^

Underlying mechanisms for developing this more lethal and resistant form of mCRPC are amplification and structural variation in the androgen receptor (*AR*) locus including its upstream enhancer. ^7-10^ Importantly, we previously showed that amplification and alteration of *AR* in plasma, including its upstream enhancer region, was associated with significantly worse survival outcomes in mCRPC patients treated with AR-directed therapy. While compelling, our prior results did not include any pre-treatment timepoints, with all patients analyzed either on- or post-treatment with AR-directed therapy, making it challenging to determine how predictive this liquid biopsy biomarker could be.

Given this major limitation of our prior study, the primary objective of our current study was to validate whether genomic alterations in the *AR* locus including the upstream enhancer could predict significantly worse survival outcomes, including in the pre-treatment setting. After validating this finding, critical for downstream clinical translation, we also sought to identify mechanisms potentially driving worse survival in these liquid biopsy biomarker-positive patients. We thus harnessed the power of cell-free DNA epigenomics and performed both fragmentomics and methylation sequencing to delve deeper into the underlying biology of lethal mCRPC. Our findings here have the potential to lead to powerfully predictive biomarkers in patients with lethal mCRPC, one of the deadliest forms of cancer in men worldwide, with mechanistic insights that could inform an entirely new class of therapeutics in the future.

## Results

### *AR*/enhancer and *PTEN* alterations are associated with lethal mCRPC

Plasma was collected from 102 mCRPC patients from two institutions, 99 of which were treated with AR-selective inhibitors (ARSIs) (**Figure 1**; **Table S1**), with a median follow-up time of 34 months (**Table S2**). Plasma samples were collected prior to ARSI initiation in 63 patients, and during treatment in 36 patients (**Table S2**). All patients had targeted hybrid-capture NGS performed on collected plasma samples using the EnhanceAR-Seq platform, which focuses on *AR*, its upstream enhancer and 84 other genes important to prostate cancer^7^ (**Table S3-S5**). EnhanceAR-Seq detected *AR*/enhancer alterations in 35% (35/99) of all analyzed plasma samples (**Figure 2A** and **Figure S1**). Further, applying EnhanceAR-Seq to pre-treatment plasma, *AR*/enhancer alterations were detected in 44% (28/63) of samples, which correlated with significantly worse progression-free survival (PFS) (HR = 2.12, *p* = 0.01) and overall survival (OS) (HR = 2.48, *p* = 0.02) (**Figure 2B** and **2C**). *AR*/enhancer alterations detected in 19% (7/36) of samples collected during ARSI were also associated with profoundly worse PFS (HR = 15.38, *p* = 0.0003) and OS (HR = 15.53, p = 0.002) (**Figure 2D** and **2E**), directly validating our previously published results.^7^ Survival outcomes were also effectively stratified when restricting our analysis to just the *AR* enhancer region or *AR* gene body (**Figure S2**).

**Figure 1.**
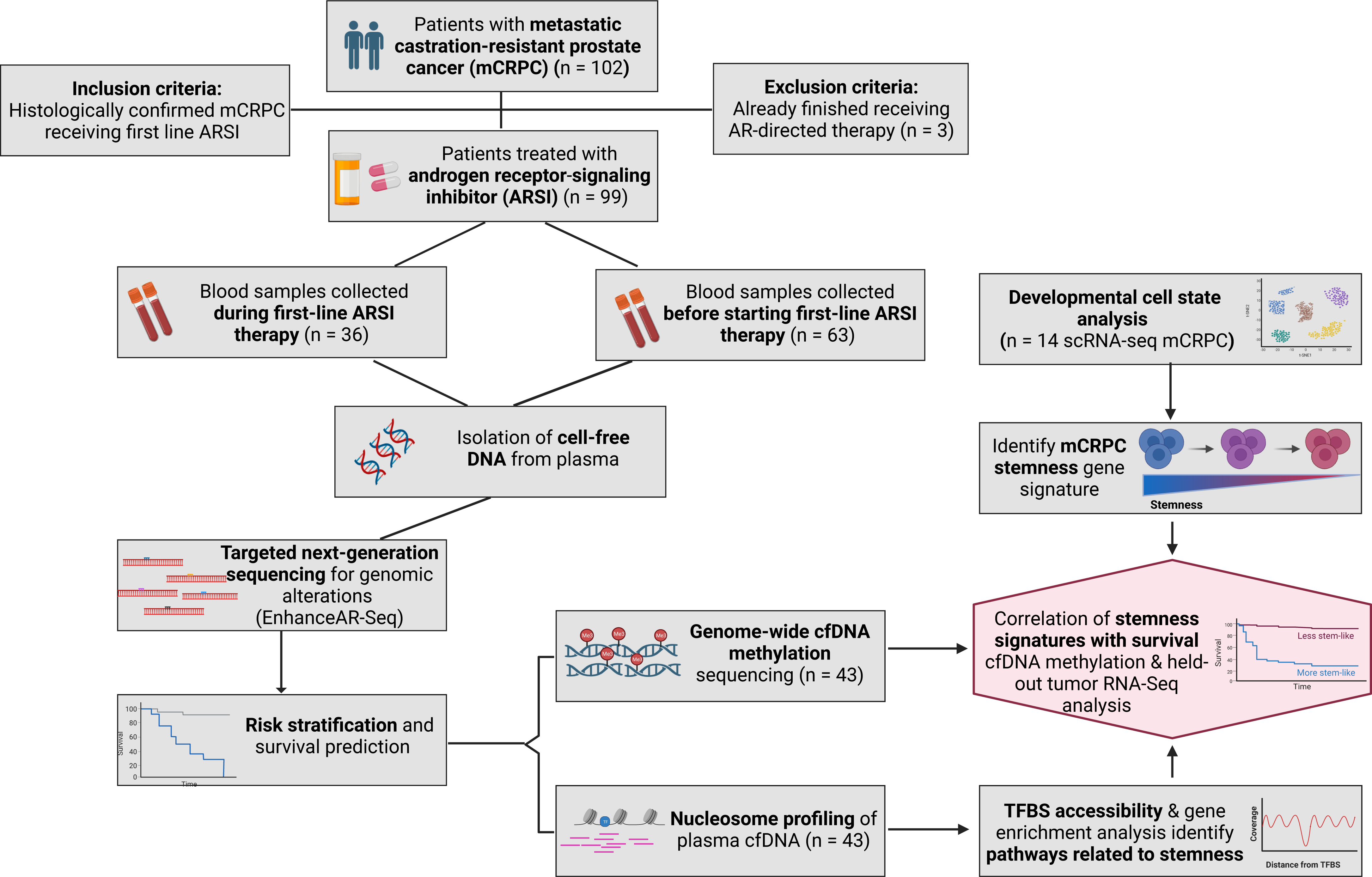
Study overview. Blood samples were collected from 102 mCRPC patients from two independent institutions, including samples collected prior to the initiation of AR-targeted therapy and during treatment. EnhanceAR-Seq applied to plasma cell-free DNA was used for the detection of genomic alterations and risk stratification of mCRPC patients. Genome-wide EM-seq for 43 patients was done using plasma collected prior to the initiation of first-line AR-targeted therapy. Nucleosome profiling of binding sites for 377 transcription factors in plasma cfDNA was done using Griffin to identify pathways enriched in AR-altered lethal mCRPC patients. scRNA- seq data from 14 separate mCRPC patients was analyzed to identify stemness markers using CytoTRACE, which were used to define a stemness signature that was validated in both plasma EM-seq and tumor tissue bulk RNA-seq data. *AR*, androgen receptor gene; EM-seq, enzymatic methylation sequencing; EnhanceAR-Seq, Enhancer and neighboring loci of Androgen Receptor Sequencing; mCRPC, metastatic castration-resistant prostate cancer; RNA-seq, RNA sequencing; scRNA-seq, single-cell RNA sequencing.

**Figure 2.**
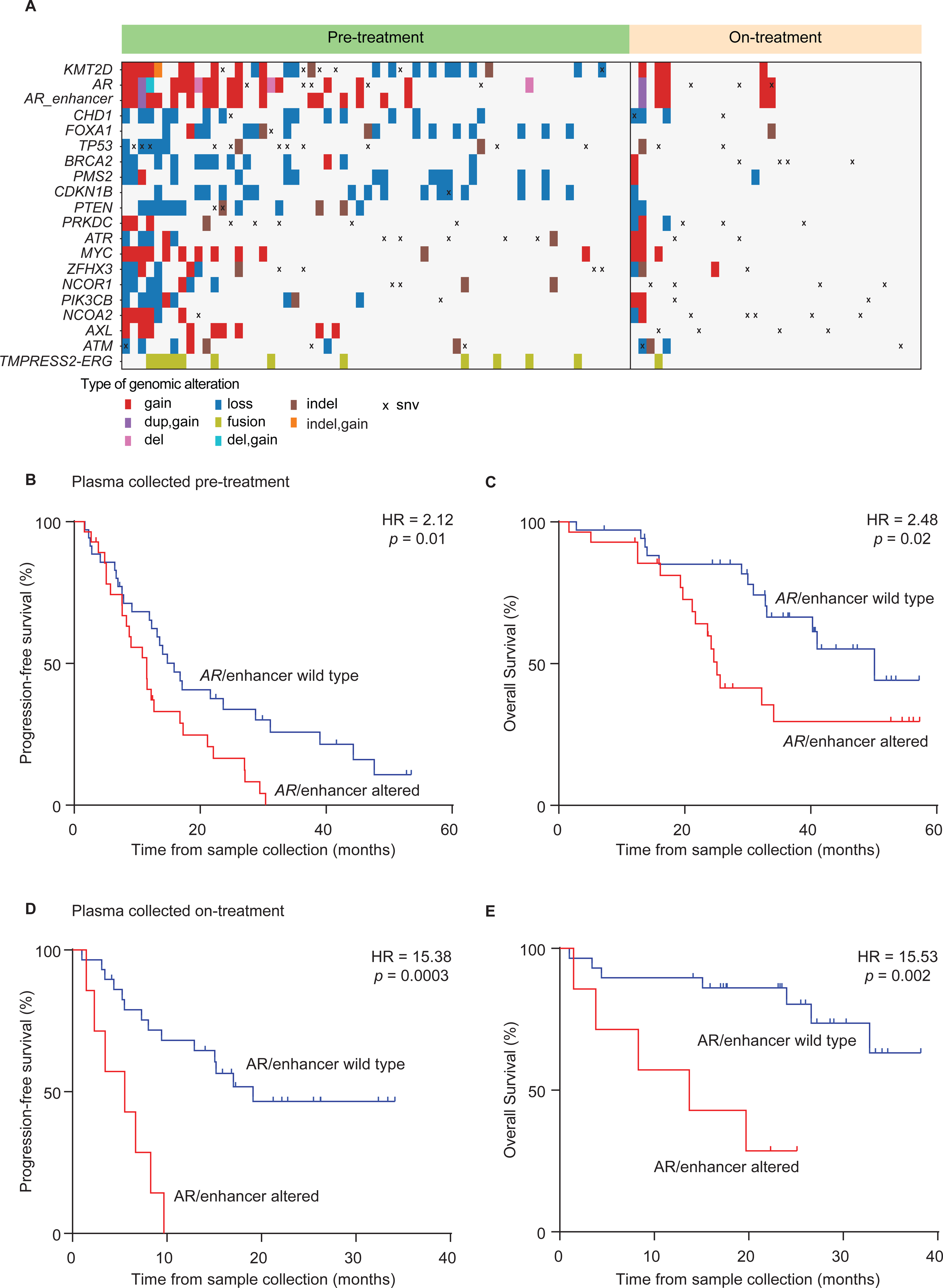
Genomic characterization of mCRPC plasma cell-free DNA. (A) Genomic alterations detected in plasma cfDNA including the androgen receptor (*AR*) and enhancer region upstream of *AR* in pre-ARSI and on-ARSI plasma collected from mCRPC patients. Progression-free and overall survival Kaplan-Meier analysis according to *AR*/enhancer alteration status in plasma collected (B-C) before starting first-line ARSIs and (D-E) during first-line ARSIs. *p* values were calculated by the log-rank test and hazard ratios (HRs) by the Mantel-Haenszel method. ARSI, androgen-receptor signaling inhibitor; cfDNA, cell-free DNA; mCRPC, metastatic castration-resistant prostate cancer.

Looking beyond *AR*, the most frequent genomic events detected in pre-treatment plasma cell-free DNA (cfDNA) were in *KMT2D* (46%) and *CHD1* (24%), consistent with previous genomic studies of mCRPC (**Figure 2A** and **Figure S1**).^7-9^ We also detected *TMPRSS2::ERG* gene fusions in the pre-treatment plasma of 10 patients (16%) (**Figure 2A** and **Figure S1**), and observed mutations in distinct biological pathways (**Figure S3**). Still, other than *AR*, we did not observe any association of these gene mutations with prognosis except for *PTEN* (**Figure S4**). Indeed, *PTEN* copy number loss was significantly associated with decreased PFS and OS (**Figure S4A** and **S4B**). This association remained significant when including cases with *PTEN* mutation (**Figure S4C** and **S4D**). Strikingly, patients with alterations in both *AR* and *PTEN* genes had even shorter PFS and OS (**Figure S4E** and **S4F**). These findings suggest that mCRPC patients with *AR* and *PTEN* alterations detected in pre-treatment plasma have more lethal mCRPC (corroborating prior tumor tissue sequencing data^11^) and could be candidates for escalated upfront therapy.

### Stemness features underpinnings of *AR*/enhancer altered lethal mCRPC

Given our previously published^7^ and newly validated findings showing that *AR*/enhancer alteration in plasma cell-free DNA is associated with significantly worse survival in mCRPC, we next queried the underlying epigenomic mechanism for this increased lethality. We thus performed genome-wide enzymatic methylation sequencing (EM-seq) on 43 patient plasma samples collected before starting ARSI treatment (**Table S6**). We began by investigating nucleosome occupancy across transcription factor (TF) binding sites genome-wide for 377 TFs obtained from the Gene Transcription Regulation Database (GTRD)).^12^ Binding sites for 52 TFs were more accessible compared to 325 being less accessible in the plasma cfDNA of *AR*/enhancer altered lethal mCRPC (**Table S7** and **S8**). For example, binding sites for the developmental regulator *HOXB13* were more accessible in *AR*/enhancer altered lethal mCRPC patients (**Figure 3A** and **3B**). Conversely, binding sites for another developmental regulator, *FOXO1,* were less accessible in *AR*/enhancer altered lethal mCRPC (**Figure 3C** and **3D**).

**Figure 3.**
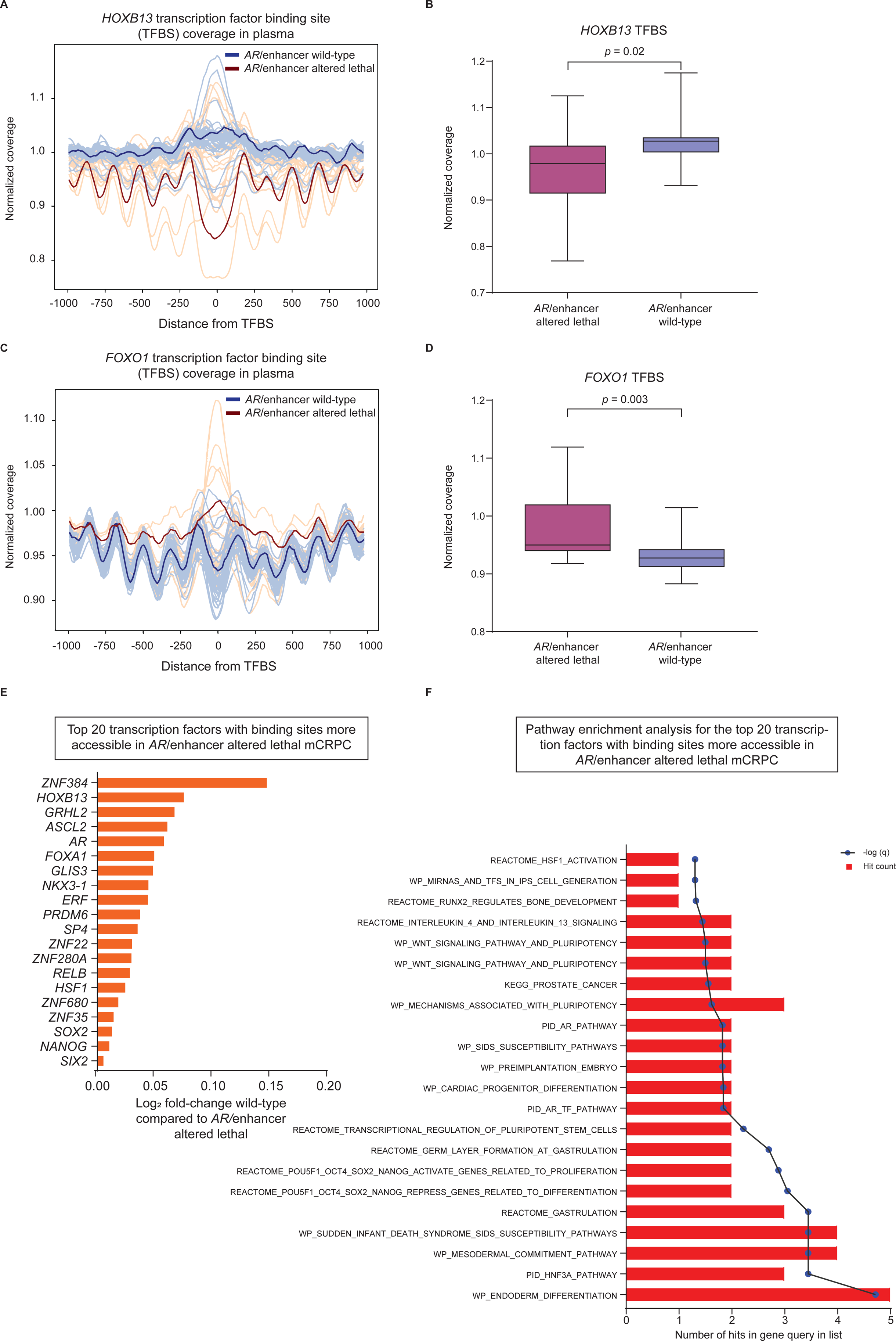
Nucleosome profiling of plasma cell-free DNA in AR/enhancer altered lethal mCRPC patients. Nucleosome profiling of 10,000 TFBSs associated with 377 transcription factors from GTRD (Methods) to infer transcription factor activity from plasma cfDNA. Central coverage profiles for transcription factors (A,B) *HOXB13* and (C,D) *FOXO1* in *AR*/enhancer altered lethal versus *AR*/enhancer wild-type mCRPC patients. Data in B,D are represented as box and whisker plots, with *p* values calculated by Student’s t test. (E) Top 20 transcription factors with binding sites found to be most accessible in *AR*/enhancer altered lethal mCRPC patients versus AR/enhancer wild-type mCRPC, and (F) gene set enrichment analysis of these 20 transcription factors with –log10(q) and hit count in the query list shown. *AR*, androgen receptor gene; cfDNA, cell-free DNA; GTRD, Gene Transcription Regulation Database; mCRPC, metastatic castration-resistant prostate cancer; TFBS, transcription factor binding site.

We were intrigued by the differential nucleosomal accessibility of these developmentally oriented transcription factors, and thus broadened our analysis. Looking across the 20 most accessible TFs in plasma cfDNA (**Figure 3E**) which we further validated in tissue using TCGA (**Figure S5**), we performed gene set enrichment analysis and strikingly observed enrichment for pathways associated with stem cell development (**Figure 3F**).

We then wondered if we could more formally query stemness in these lethal mCRPC patients using cfDNA. We hypothesized that we might be able to infer stemness genomically, in a manner analogous to the gene counts-based methodology (CytoTRACE) applied to single-cell RNA sequencing data by Gulati et al.^13^ (**Figure 4A**). We thus began by applying CytoTRACE to publicly available single-cell RNA-seq data from 14 mCRPC patients.^14^ The prostate adenocarcinoma cluster of cells (n = 835) was identified in this dataset based on expression of the *AR* and *KLK3* genes (**Figure 4B**). Using CytoTRACE, we then obtained a developmental stemness score for each adenocarcinoma tumor cell within this dataset, with cells having a high CytoTRACE score being more stem-like, and those with a low score being less stem-like.^13^ Next we identified the genes most differentially expressed in these more stem-like versus less stem-like mCRPC adenocarcinoma cells (**Figure 4C**). Corroborating expected biology, these included genes known to play major roles in the processes associated with stemness, cellular proliferation, and self-renewal.^15-19^ Further, suggesting a connection with *AR*/enhancer altered lethal mCRPC, the ten genes most associated with stemness were promoter hypo-methylated in this more lethal disease state, while the ten genes least associated with stemness were promoter hyper-methylated (**Figure 4D and 4E**).

**Figure 4.**
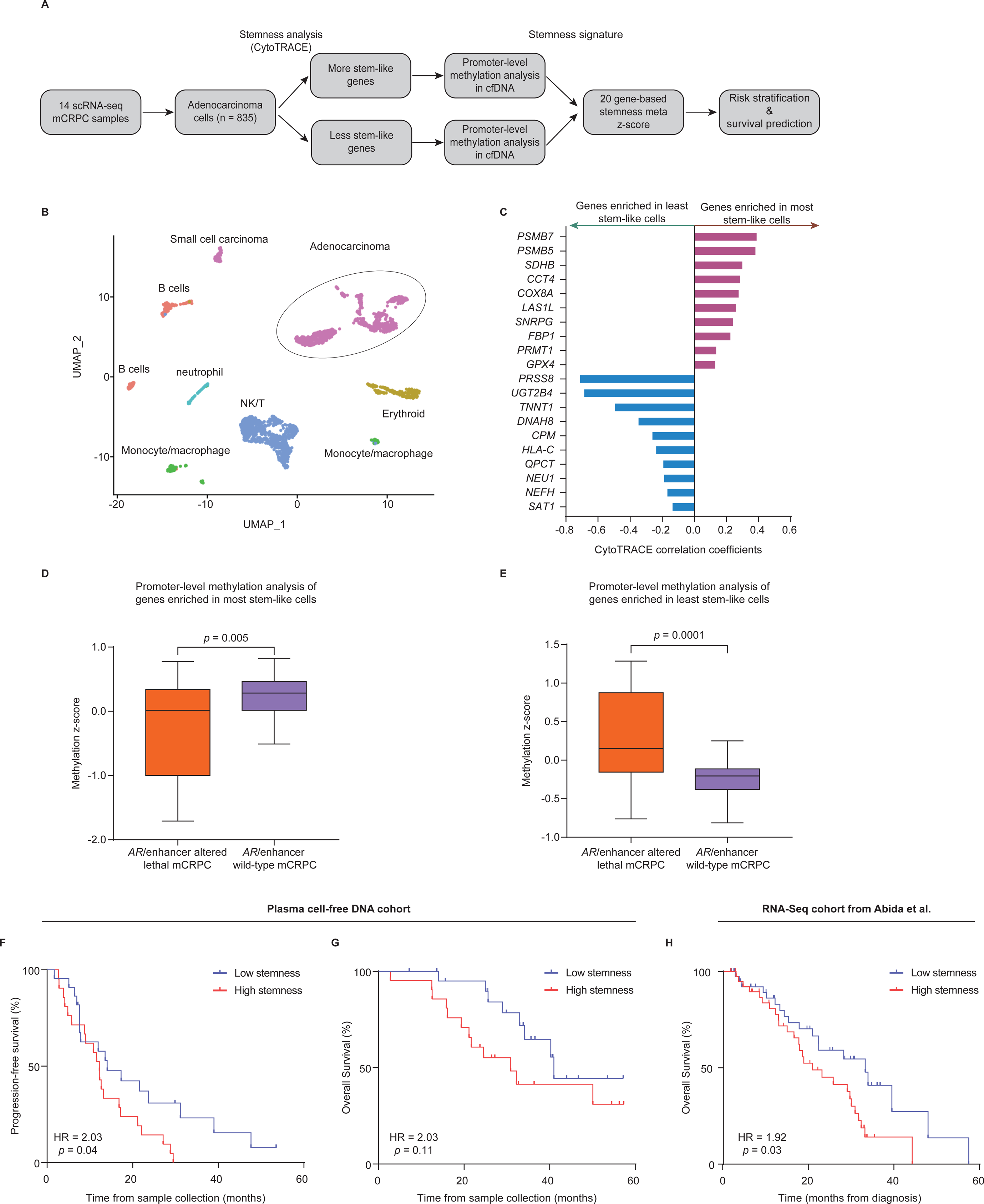
Prognostic stemness signatures associated with lethal mCRPC. (A) Schema describing the stemness analysis workflow. (B) UMAP decomposition of scRNA-seq data from 14 mCRPC patients with the adenocarcinoma cluster comprised of 835 cells highlighted. (C) Top 10 differentially expressed genes in most stem-like cells and least stem-like cells after single-cell analysis of mCRPC adenocarcinoma cells by CytoTRACE (Methods). Comparison of promoter-level methylation of the (D) most and (E) least stem-like genes with mCRPC type after metagene analysis (Methods). Data are shown as box and whisker plots, with *p* values calculated by Student’s t test. (F-G) Stemness measured from plasma EM-seq (Methods) correlated with worse survival outcomes by Kaplan-Meier analysis. (H) Validation of the stemness signature in an external cohort of 80 mCRPC patients with tumor tissue profiled by bulk RNA-seq. For Kaplan-Meier analyses, *p* values were calculated by the log-rank test and hazard ratios (HRs) by the Mantel-Haenszel method. EM-seq, enzymatic methylation sequencing; mCRPC, metastatic castration-resistant prostate cancer; RNA-seq, RNA sequencing; scRNA-seq, single-cell RNA sequencing; TFBS, transcription factor binding site; UMAP, Uniform Manifold Approximation and Projection.

### Stemness, inferred from cell-free DNA, predicts worse survival outcomes in mCRPC

Having identified a stemness signature from external single-cell RNA sequencing data and connecting it to *AR*/altered lethal mCRPC, we next wondered if this stemness signature was also prognostic. We thus queried pre-treatment plasma cfDNA, and found that stemness-enriched mCRPC patients had worse survival outcomes (**Figure 4F and 4G**) including in multivariate analysis with eight covariates that included ctDNA and PSA levels (**Table S9**). We next, importantly, applied our stemness signature to a completely held-out validation cohort of 80 mCRPC patients profiled by tumor tissue RNA-seq.^20^ Strikingly, despite differences in cohorts, sequencing substrate and sequencing modality, the 20-gene stemness signature again validated prognostically, with stemness-high patients again having significantly worse survival than stemness-low patients (**Figure 4H**).

## Discussion

Despite the clinical success of second-generation AR-directed therapies such as abiraterone and enzalutamide in mCRPC, approximately one-third of patients invariably develop lethal resistant disease^2,5^. In this study, we first validated our previously published finding that *AR*/enhancer alterations in cell-free DNA are highly prognostic,^7^ extending upon our earlier findings by further showing that pre-treatment *AR*/enhancer alterations are also strikingly prognostic. We further learned that *PTEN* alterations measured from cell-free DNA are associated with worse survival outcomes, consistent with the published tumor tissue sequencing literature^11,21-24^.

While others have also observed alterations in *AR*/enhancer associating with lethal disease^8,10^, it remains unclear what the underlying mechanism for this increased lethality is. To query this practically, we performed genome-wide cell-free DNA EM-seq to learn the epigenomic differences between *AR*/enhancer altered lethal disease and its less deadly *AR*-wild-type counterpart. We learned that not only were there stark epigenomic differences between these mCRPC subtypes, but there appeared to be an underlying pattern based on stemness pathways. Delving more deeply using externally available RNA sequencing data, we learned and validated a stem cell signature, which was also enriched in our own cell-free DNA data from *AR*/enhancer altered lethal mCRPC.

Indeed, it has been shown that stemness features are associated with cancer aggressiveness, therapeutic resistance, and increased lethality.^13,25-27^ Gulati and colleagues used technology analogous to ours, applied to breast cancer, and observed an aggressive cancer stem cell state, with genomic markers that could be selectively inhibited to induce tumor cytotoxicity.^13^ Recently, we also applied similar technology to pancreatic cancer, and identified a stem-like state there that was associated with significantly worse survival outcomes.^27^ Despite these prior studies in other cancer types, the work here in mCRPC is novel, as we were able to validate our stem-like signature using cell-free DNA data, while the prior works relied on tumor tissue RNA sequencing data for validation.

We acknowledge certain limitations of our study. First, we included patient samples from two separate institutions which could introduce unintended biases. Nevertheless, we see this as more of a strength than a weakness, as it suggests that our results are more likely to further validate in a broader multi-institutional setting. Second, while our risk stratification was based on *AR*/enhancer locus alterations, other genomic alterations involving *PTEN*, *RB1*, *TMPRSS2*::*ERG* and DNA damage repair genes could have been used for risk stratification as well. Indeed, while we show data here that mCRPC patients with *PTEN* alterations in cell-free DNA also had significantly worse outcomes, we wanted to remain rather focused on *AR*/enhancer altered versus wild-type mCRPC, to cleanly validate our previously published results^7^ while delving more deeply into underpinning mechanisms. Third, the stemness signature genes obtained in our study are not purely canonical in nature, for example it does not include the previously published Yamanaka factors.^28^ Nevertheless, we identified this stemness signature in a highly objective fashion using completely external data, and it includes genes shown to be important in development, cellular proliferation, and epithelial-mesenchymal transition.^15-19^

In conclusion, our study provides further evidence that genomic alterations in the *AR* locus including the upstream enhancer region are remarkably prognostic, with pre-treatment predictive potential. Delving deeper into the underlying mechanism, we identify cancer stemness as a lethality factor that could explain why these *AR*/enhancer altered patients have such worse survival outcomes. Interestingly, we were able to infer cancer stemness from the cell-free DNA liquid biopsy results themselves, representing a significant methodological advancement for the field in addition to the important biological implications.

## Data Availability

All data produced in the present study are available upon reasonable request to the authors

## Acknowledgments

We are grateful to the patients, and their families, involved in this study. Images from Biorender.com were used to create **Figure 1**. This work was supported by NIH under the award F31 CA265010-01 (**J.W.**). This work was also supported by the National Cancer Institute (NCI) under award number K08CA238711 (**A.A.C.**), and the National Center for Advancing Translational Sciences (NCATS) under award number UL1TR002345 (Principal Investigator, Bradley Evanoff; **A.A.C.**). This work was additionally supported by LEAP competition through the Skandalaris Center for Interdisciplinary Innovation and Entrepreneurship, the Institute for Clinical and Translational Sciences, and the Siteman Cancer Center of Washington University in St. Louis under award number (Award #: 1077, **A.A.C.**), the Alvin J. Siteman Cancer Research Fund (**A.A.C.**), the Cancer Research Foundation Young Investigator Award (**A.A.C.**), and the V Foundation V Scholar Award (**A.A.C.**). The funders had no role in study design, data collection and analysis, decision to publish, or preparation of the manuscript.

## Author Contributions

Chauhan and Alahi contributed equally to this work. Dr. Chaudhuri had full access to all data in the study and takes responsibility for the integrity of the data and the accuracy of the data analysis. Concept and design: Oliver, Pachynski, Maher and Chaudhuri.

Acquisition, analysis, or interpretation of data: All authors.

Drafting of the manuscript: Chauhan, Alahi, Sinha, Mueller, Webster, Harris, and Chaudhuri. Critical revision of the manuscript for important intellectual content: All authors.

Statistical analysis: Alahi, Sinha, Chauhan, Webster and Dang. Obtained funding: Oliver, Pachynski, Maher and Chaudhuri.

Administrative, technical, or material support: Qaium, Oliver, Pachynski, Maher and Chaudhuri.

Supervision: Oliver, Pachynski, Maher and Chaudhuri.

## Methods

### Study design and patient enrollment

In this multi-institutional study, plasma samples from 102 mCRPC patients were collected from two independent institutions (n = 55 from Tulane, n = 47 from WashU). Plasma samples were collected prior to the initiation of first-line AR-targeted therapy (n = 63) and during treatment (n = 36). The WashU cohort, comprised of 47 patient samples, was prospectively enrolled between February 2019 and September 2021. Of the 47 patients enrolled, 19 received abiraterone and 28 received enzalutamide. The Tulane cohort, comprised of 55 patient samples, was collected between March 2015 and January 2020. Three patients from the Tulane cohort were excluded, as they had already received AR-directed therapy and samples were collected post-treatment. Of the remaining 52 patients, 46 received abiraterone and 6 received enzalutamide. All patients were maintained on standard androgen deprivation therapy (i.e., luteinizing hormone-releasing hormone receptor agonist or antagonist). Prior treatment with other systemic agents, including chemotherapy, was allowed. Patients with evidence of any active non-prostate malignancy other than localized skin cancer were excluded from the study. All samples were collected with informed consent and institutional review board approval (IRB) (Washington University IRB # 201411135 and Tulane IRB # 992885) in accordance with the Declaration of Helsinki. Sample IDs in the manuscript or in supplemental data are not known to anyone outside of the research group.

### Sample collection and processing

For the WashU cohort, peripheral blood samples between 10 and 20 ml were collected from each patient in K2EDTA vacutainer tubes (Becton Dickinson, Franklin Lakes, NJ) at the time of study enrollment. Plasma was separated by centrifugation at 1,200g for 10 minutes at room temperature. Plasma-depleted whole blood (PDWB) was collected and frozen at −80°C for isolation of germline DNA. Plasma was collected, followed by another spin at 1,800g for 5 minutes. As previously described, both plasma and PDWB were frozen at −80°C before the isolation of cfDNA and germline gDNA, respectively. Double-spun plasma and PDWB were similarly obtained from patients in the Tulane cohort.

### Nucleic acid isolation

Circulating cfDNA was isolated from plasma using the QIAamp Circulating Nucleic Acid Kit (Qiagen, Hilden, Germany) according to the manufacturer’s instructions. Germline DNA was extracted from PDWB using the QIAamp DNA Blood Mini Kit (Qiagen). DNA was then quantified by the Qubit dsDNA High Sensitivity Assay (Thermo Fisher Scientific, Waltham, Massachusetts) and quality was further assessed using an Agilent 2100 Bioanalyzer (Agilent Technologies, Santa Clara, California).

### Enhancer and neighboring loci of Androgen Receptor sequencing

**(EnhanceAR-Seq)** EnhanceAR-Seq was performed on plasma cfDNA along with matched germline DNA as previously described.^7^ Briefly, a median of 32 ng of plasma cfDNA was input into sequencing library preparation based on the percentage of cfDNA in the 70-450 bp region of the Bioanalyzer electropherogram. Corresponding germline DNA was fragmented to ∼180 bp size fragments prior to library preparation using a LE220 focused ultrasonicator (Covaris, Woburn, Massachusetts). 32 ng of sheared germline DNA along with unsheared plasma cfDNA was used for library preparation using the KAPA HyperPrep kit (Roche, Basel, Switzerland) with barcoded adapters containing demultiplexing, deduplicating and duplexing unique molecular identifiers. Targeted hybrid capture was performed per the standard CAPP-Seq method.^29-31^ We used a focused gene panel to target the complete *AR* gene body (including introns), 30 kb of the *AR* enhancer, and exons of 84 other genes that have been shown to harbor genomic alterations in mCRPC.^8^ EnhanceAR-Seq libraries were then sequenced, as previously described,^7^ on an Illumina HiSeq4000 with 2×150 bp paired-end reads with 12 samples sequenced per lane, dedicating approximately 50 million total reads per sample.

### cfDNA sequencing data pre-processing

Cell-free DNA sequencing reads were mapped to the hg19 human reference genome using bwa mem. Fgbio was then used to group reads by unique molecular identifiers, requiring a minimum mapping quality score of 10. Consensus reads were called, requiring a base quality Phred score > 30. Consensus reads were then re-mapped using bwa mem for downstream analysis (including structural variation, copy number alteration and single nucleotide variant calling).

### Structural variation (SV), copy number alteration (CNA) and small mutation analysis

We used PACT, a standardized ctDNA pipeline for detection of SVs, CNAs and small mutations recently published by our group.^32^ Plasma cfDNA structural variant calling was performed using the default settings of the PACT workflow. Briefly, SV candidates were called using an ensemble of SV callers, including Delly^33^, Lumpy^34^ and Manta^35^, and consensus calls were then filtered based on their overlap with regions included in the targeted panel and by excluding SVs that overlapped with blacklisted or low complexity regions. Sequencing/alignment artifacts and possible germline events were then removed by filtering out any SV with read support in matched controls (PDWB) or in a panel of samples from 24 healthy individuals. Finally, ≥1 split read and ≥1 supporting discordant paired-end read were required. SVs that passed these filters were then annotated using snpEff.^36^

Copy number alteration (CNA) calling was also performed using the default settings of the PACT workflow. First the log-transformed ratio of sequencing depth between cfDNA and matched control (PDWB) samples was calculated, while correcting for repeat content and GC content biases. Then, recentralization of log depth ratios was performed using copy number control regions to account for depth biases resulting as an artifact of targeted sequencing. This task was accomplished using the recommended CNVkit workflow.^37^ Finally, regions with a log depth ratio deviating from the control regions by >3 standard deviations from the mean were considered CNAs.

For single nucleotide variant (SNV) calling, we again used the default settings of the PACT workflow. The workflow began by creating a list of candidate SNVs using Mutect^38^, Strelka^39^, VarScan^40^ and Pindel^41^. Additionally, samples were genotyped using GATK’s HaplotypeCaller^42^ and the DoCM database v3.2.^43^ Results from the four SNV callers and from HaplotypeCaller were then combined and decomposed using Vt’s decompose function.^44^ Read counts were re-calculated using bam-readcount^45^ and filtering was performed based on frequency in gnomAD (<0.1% population frequency), mapping quality (<15% reads with mapq0), read depth (>8), read support (>2) and allele frequency (>0.1%). To address the false positives likely to occur due to deep sequencing and low expected allele frequencies, background error suppression was performed by genotyping remaining variants in our panel of healthy normal individuals (n = 24) using HaplotypeCaller. Any event with read support in >2 healthy individuals was removed. SNVs were annotated using vep v100.^46^

### Enzymatic Methylation sequencing (EM-Seq) library preparation

EM-seq libraries were prepared from plasma cfDNA collected prior to AR-targeted therapy in 43 mCRPC patients using the NEB Next Enzymatic Methyl-seq kit (New England Biolabs, Ipswitch, Massachusetts) following the manufacturer’s instructions. Briefly, 10ng of plasma cfDNA was mixed with 1% (0.1ng) of unmethylated lambda DNA (used as spike-in for quality control). EM-seq libraries were then sequenced on a NovaSeq 6000 sequencer (Illumina, San Diego, California) in paired-end 150 bp mode with a median of ∼16x genome-wide read coverage. Sequencing metrics are provided in Table S6.

### Alignment of methylation samples

We used BISCUIT for EM-seq next-generation sequencing alignment and methylation calling. After trimming adapters using flexbar^47^, paired Fastq files were provided to the BISCUIT align sub command with hg38 human genome as the reference. After alignment, BISCUIT pileup was used to compute cytosine retention with base quality ≥ 20 and mapping quality ≥ 40. The outputted vcf file was then converted to a bed file using BISCUIT vcf2bed.

### EM-seq bioinformatic quality control

To check the quality of the aligned samples, we primarily used samtools-flagstat.^48^ Samtools-flagstat provides several important quality-control metrics such as total reads, duplicate reads, and mapping percentage.^48^ By generating histograms, we further checked fragment length distributions to confirm that cell-free DNA sample sequencing was in the expected nucleosomal distribution. Using the samtools depth command, we found the median coverage across plasma cell-free DNA samples was 16x (which is what we aimed for).

To confirm the completeness of enzymatic methylation conversion for our EM-seq libraries, we utilized unmethylated control phage DNA that was included in each sample. We first used the BISCUIT align subcommand on the post-conversion paired Fastq files for each patient with the Enterobacteria phage lambda genome as the reference sequence. Duplicate reads were then marked using dupsifter. The resulting output SAM files were then converted to BAM files and subsequently indexed using samtools. BISCUIT pileup was then used to calculate cytosine retention with base quality ≥ 20 and mapping quality ≥ 40 for each BAM file, outputting a TSV file containing the average methylation beta values at each CG, CHG, CHH, and CH site for every sample. The average CG methylation beta values were then extracted from each VCF file and subtracted from 1 to obtain the conversion rate, which was >98% across the full cohort (**Table S6**).

### Cell-free DNA nucleosomal profiling

Here we adapted methodology of analyzing the cfDNA fragmentation pattern from which nucleosomes occupancy can be inferred using methylation data.^49,50^ Leveraging the Griffin framework^51^, we performed nucleosome profiling analysis on methylation bam files. Adhering to Griffin’s guidelines, we first delineated the mappable regions for each sample, while also addressing potential GC bias. To this end, we utilized the Umap multi-read mappability tracks sourced from the UCSC genome browser, complemented by Griffin’s dedicated wrapper, to pinpoint the mappable genomic regions. Recognizing the unique GC bias inherent to each sample, especially when working with cfDNA where fragment lengths can vary, we employed Griffin’s ‘fragment length model’ based implementation for the requisite GC correction.

We then delved into TF binding site analysis, employing 377 TFs with about 10,000 sites from the Gene Transcription Regulation Database (GTRD)^12^ as recommended.^51^ We selected central coverage as our primary output feature, consistent with published Griffin analyses due to its high sensitivity. For detailed Griffin execution, refer to https://github.com/adoebley/Griffin/wiki.

### Differential expression analysis of TFs in blood and localized prostate cancer patients

We also employed RNA expression in tumor tissue and peripheral blood as an orthogonal validation method to cell-free DNA nucleosomal analysis. Using the UCSC Xena online tool^52^, we aimed to compare TF expression between localized prostate cancer and blood cells. Within this browser-based tool, we engaged with the ‘main category’ and ‘study’ options. Specifically, for the main category, we selected the differential gene expression analysis between TCGA_Prostate_Adenocarcinoma (n = 496) and GTEX_Blood (N = 337). Subsequently, the most versus least accessible TFs that we learned in mCRPC cell-free DNA was verified in TCGA_Prostate_Adenocarcinoma versus GTEX_Blood using the Student’s t test (**Figure S5**).

### Gene set enrichment analysis of the top 20 most accessible TFs

Gene set enrichment analysis was performed with ToppFun^53^ using the 20 most accessible TFs in *AR*/enhancer altered lethal mCRPC cell-free DNA. Statistical significance was assessed with the Benjamini-Hochberg procedure, using an adjusted p-value cutoff of 0.05 (**Figure 3F**).

### Stemness analysis in tumor tissue and plasma

Tumor tissue single-cell RNA-seq data from fourteen lethal prostate cancer patients were UMAP decomposed from He et al.^14^ We used *AR* and *KLK3* gene expression to identify the dominant prostate adenocarcinoma cluster of 835 cells. There was a separate small cell carcinoma cluster of cells that was identified based on *CHGA* upregulation, and not further considered. Next, we log-transformed the prostate adenocarcinoma single-cell RNA-seq gene expression data, filtered out ribosomal genes and kept only the protein-coding genes. We then ran CytoTRACE^13^ to identify the genes most associated with stemness in the data. We took the top 500 most stem-like and 500 least stem-like genes from this step and denoted these gene lists as Smore and Sless, respectively.

Next, we calculated promoter methylation levels in the cell-free DNA EM-seq cohort by averaging CpG methylation levels in the promoter regions of Smore and Sless. We ranked these promoter methylation scores by variance and denoted this ranked list as M. Given the lists Smore and M, we took all the genes between the 75^th^ and 98^th^ percentiles of each, and denoted these as S’more and M’. Finally, we took the top 10 stem-associated genes from S’more ∩ M’ and called these signature genes SIGmore. Similarly, with Sless, we took the top 10 genes from S’less ∩ M’ and called them SIGless.

To apply SIGmore to the pre-treatment cfDNA EM-seq data and stratify survival, we took the z-score of each gene corresponding to SIGmore and then created a metagene by combining them. We used a similar approach to stratify survival using SIGless. We then combined SIGmore and SIGless into SIGcombined. This was done by inverting the methylation value of SIGless and then taking the z-score and creating a metagene by combining with SIGmore (**Figure 4F and 4G**). We then median-split the metagene data to make it categorical prior to Kaplan-Meier survival analysis.

To further validate our stemness metagene in an external cohort, we used mCRPC tumor tissue bulk RNA-seq data from cBioPortal with previously calculated z-scores (prad_su2c_2019).^20^ Within this cohort, 80 patients were documented with overall survival data. Again, we inverted the z-score of the SIGless genes by multiplying by −1, and combined with SIGmore to yield the RNA-seq equivalent of SIGcombined. We again median-split the metagene data to make it categorical prior to Kaplan-Meier survival analysis (**Figure 4H**).

### Statistical analysis

ToppFun^53^ was used for gene set enrichment analysis of the most accessible TFs discovered in the plasma of *AR*/enhancer altered lethal mCRPC patients. For combining methylation levels across gene promoters or expression levels across genes, z-score normalization was used. All two-group statistical analyses were conducted using the Student’s t test. The Benjamini Hochberg test was performed when multiple hypotheses testing was conducted. We used a 0.05 significance threshold for all statistical tests employed. Survival plots were generated using Kaplan–Meier analysis, and the log-rank test was used to determine statistical significance, and Mantel-Haenszel method for calculating hazard ratios. Prism 9 (GraphPad Software, San Diego, California) was used for all clinical-correlative statistical and survival analyses, except for multivariate cox proportional hazards model analyses which were performed using the lifelines python package.

## Supplemental Information

### Supplementary figures

**Figure S1.**
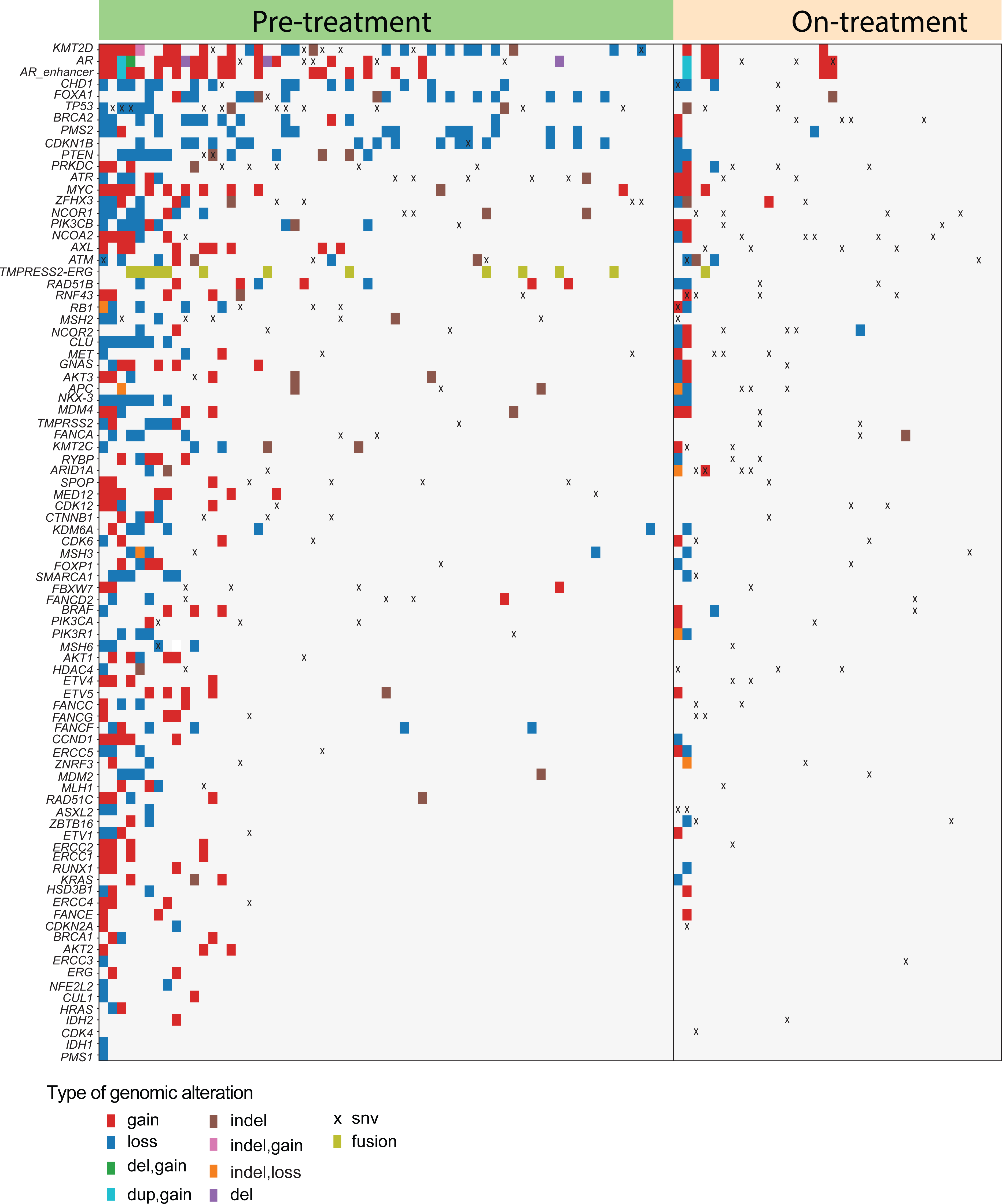
Genomic landscape of metastatic castration-resistant prostate cancer (mCRPC) samples profiled using EnhanceAR-seq. EnhanceAR-Seq, Enhancer and neighboring loci of Androgen Receptor Sequencing.

**Figure S2.**
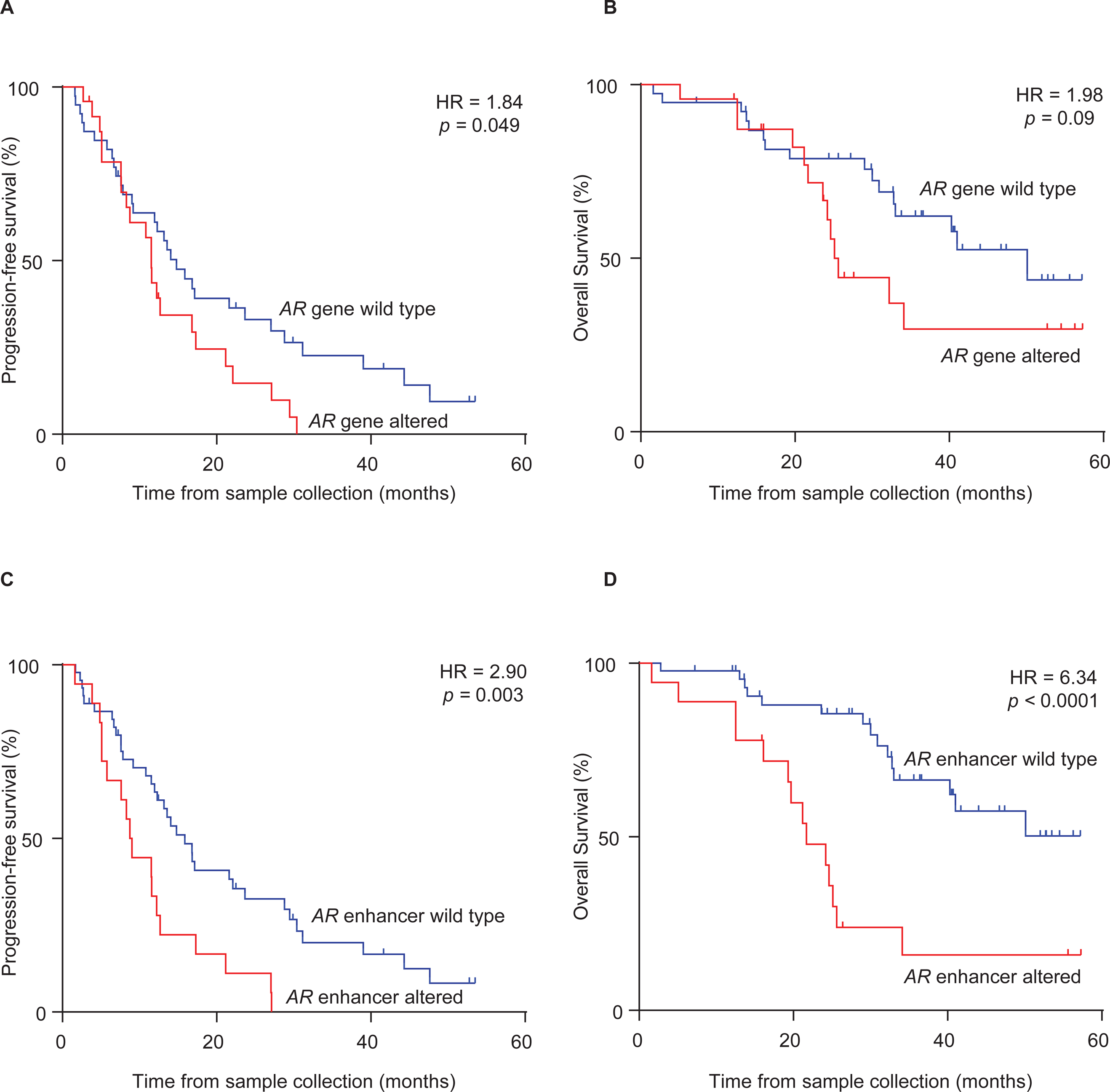
Kaplan-Meier analysis based on plasma collected prior to first-line androgen receptor-signaling inhibitor (ARSI) treatment according to (A,B) Androgen receptor (AR) gene body status and (C,D) AR enhancer region status in 63 patients with metastatic castration-resistant prostate cancer (mCRPC). *p* values were calculated by the log-rank test and hazard ratios (HRs) by the Mantel-Haenszel method.

**Figure S3.**
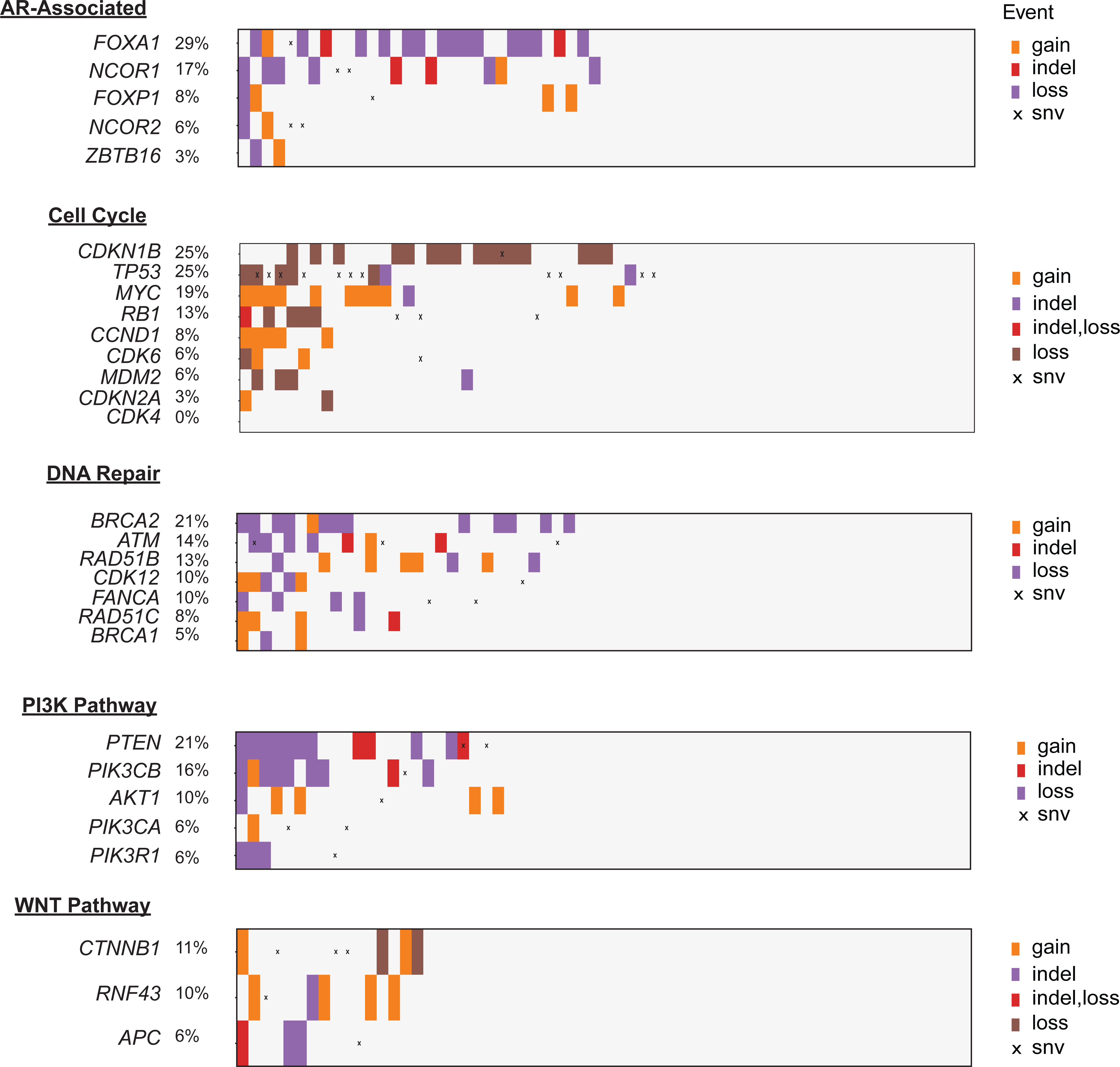
Summary of alterations in biologically relevant pathways found in pre-ARSI plasma cell-free DNA of 63 metastatic castration-resistant prostate cancer patients.

**Figure S4.**
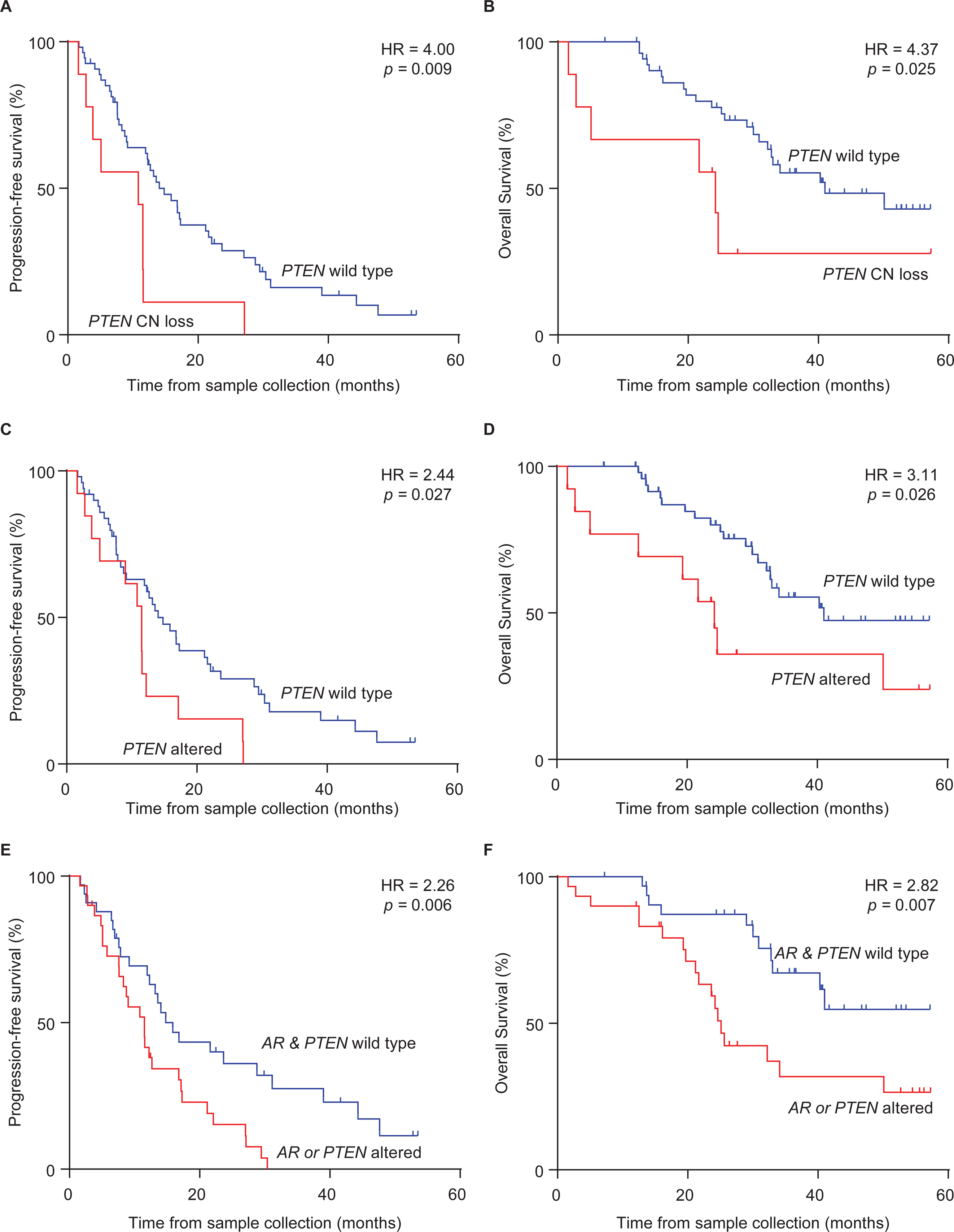
Kaplan-Meier analysis based on plasma collected prior to first-line androgen receptor-signaling inhibitor (ARSI) treatment in 63 mCRPC patients according to (A,B) *PTEN* copy number variation, (C,D) *PTEN* copy number variation or mutation, and (E,F) alteration in *AR*/Enhancer or *PTEN*. *p* values were calculated by the log-rank test and hazard ratios (HRs) by the Mantel-Haenszel method.

**Figure S5.**
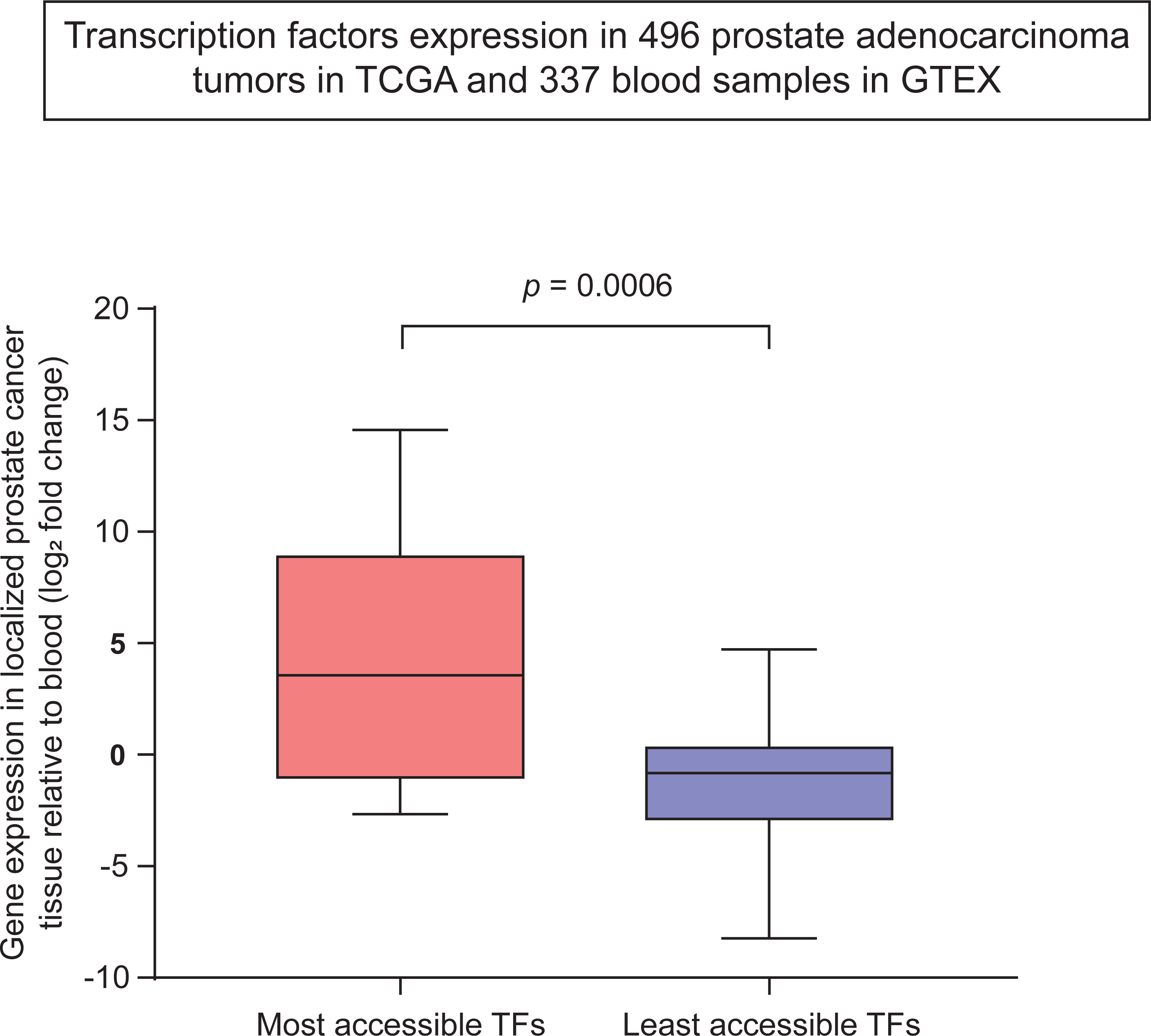
Transcription factor analysis in localized prostate cancer versus blood. Log2 fold change of the 20 TFs with most accessible binding sites in plasma cell-free DNA (see Fig. 3E) in localized prostate adenocarcinoma tumors from TCGA (n = 496) versus blood samples from GTEX (n = 337) (Methods). *p* value was calculated by Student’s t test. GTEX, The Genotype-Tissue Expression project; TCGA, The Cancer Atlas Genome Atlas project; TF, transcription factor.

## References

1. Beer, T.M., Armstrong, A.J., Rathkopf, D.E., Loriot, Y., Sternberg, C.N., Higano, C.S., Iversen, P., Bhattacharya, S., Carles, J., Chowdhury, S., et al. (2014). Enzalutamide in metastatic prostate cancer before chemotherapy. N Engl J Med 371, 424–433. 10.1056/NEJMoa1405095.

2. Scher, H.I., Fizazi, K., Saad, F., Taplin, M.E., Sternberg, C.N., Miller, K., de Wit, R., Mulders, P., Chi, K.N., Shore, N.D., et al. (2012). Increased survival with enzalutamide in prostate cancer after chemotherapy. N Engl J Med 367, 1187–1197. 10.1056/NEJMoa1207506.

3. Fizazi, K., Scher, H.I., Molina, A., Logothetis, C.J., Chi, K.N., Jones, R.J., Staffurth, J.N., North, S., Vogelzang, N.J., Saad, F., et al. (2012). Abiraterone acetate for treatment of metastatic castration-resistant prostate cancer: final overall survival analysis of the COU-AA-301 randomised, double-blind, placebo-controlled phase 3 study. Lancet Oncol 13, 983–992. 10.1016/S1470-2045(12)70379-0.

4. Ryan, C.J., Smith, M.R., Fizazi, K., Saad, F., Mulders, P.F., Sternberg, C.N., Miller, K., Logothetis, C.J., Shore, N.D., Small, E.J., et al. (2015). Abiraterone acetate plus prednisone versus placebo plus prednisone in chemotherapy-naive men with metastatic castration-resistant prostate cancer (COU-AA-302): final overall survival analysis of a randomised, double-blind, placebo-controlled phase 3 study. Lancet Oncol 16, 152–160. 10.1016/S1470-2045(14)71205-7.

5. Antonarakis, E.S., Lu, C., Wang, H., Luber, B., Nakazawa, M., Roeser, J.C., Chen, Y., Mohammad, T.A., Chen, Y., Fedor, H.L., et al. (2014). AR-V7 and resistance to enzalutamide and abiraterone in prostate cancer. N Engl J Med 371, 1028–1038. 10.1056/NEJMoa1315815.

6. Scher, H.I., Lu, D., Schreiber, N.A., Louw, J., Graf, R.P., Vargas, H.A., Johnson, A., Jendrisak, A., Bambury, R., Danila, D., et al. (2016). Association of AR-V7 on Circulating Tumor Cells as a Treatment-Specific Biomarker With Outcomes and Survival in Castration-Resistant Prostate Cancer. JAMA Oncol 2, 1441–1449. 10.1001/jamaoncol.2016.1828.

7. Dang, H.X., Chauhan, P.S., Ellis, H., Feng, W., Harris, P.K., Smith, G., Qiao, M., Dienstbach, K., Beck, R., Atkocius, A., et al. (2020). Cell-free DNA alterations in the AR enhancer and locus predict resistance to AR-directed therapy in patients with metastatic prostate cancer. JCO Precis Oncol 4, 680–713. 10.1200/po.20.00047.

8. Quigley, D.A., Dang, H.X., Zhao, S.G., Lloyd, P., Aggarwal, R., Alumkal, J.J., Foye, A., Kothari, V., Perry, M.D., Bailey, A.M., et al. (2018). Genomic Hallmarks and Structural Variation in Metastatic Prostate Cancer. Cell 174, 758–769 e759. 10.1016/j.cell.2018.06.039.

9. Robinson, D., Van Allen, E.M., Wu, Y.M., Schultz, N., Lonigro, R.J., Mosquera, J.M., Montgomery, B., Taplin, M.E., Pritchard, C.C., Attard, G., et al. (2015). Integrative clinical genomics of advanced prostate cancer. Cell 161, 1215–1228. 10.1016/j.cell.2015.05.001.

10. Viswanathan, S.R., Ha, G., Hoff, A.M., Wala, J.A., Carrot-Zhang, J., Whelan, C.W., Haradhvala, N.J., Freeman, S.S., Reed, S.C., Rhoades, J., et al. (2018). Structural Alterations Driving Castration-Resistant Prostate Cancer Revealed by Linked-Read Genome Sequencing. Cell 174, 433–447 e419. 10.1016/j.cell.2018.05.036.

11. Hamid, A.A., Gray, K.P., Shaw, G., MacConaill, L.E., Evan, C., Bernard, B., Loda, M., Corcoran, N.M., Van Allen, E.M., Choudhury, A.D., and Sweeney, C.J. (2019). Compound Genomic Alterations of TP53, PTEN, and RB1 Tumor Suppressors in Localized and Metastatic Prostate Cancer. Eur Urol 76, 89–97. 10.1016/j.eururo.2018.11.045.

12. Yevshin, I., Sharipov, R., Kolmykov, S., Kondrakhin, Y., and Kolpakov, F. (2019). GTRD: a database on gene transcription regulation-2019 update. Nucleic Acids Res 47, D100–D105. 10.1093/nar/gky1128.

13. Gulati, G.S., Sikandar, S.S., Wesche, D.J., Manjunath, A., Bharadwaj, A., Berger, M.J., Ilagan, F., Kuo, A.H., Hsieh, R.W., Cai, S., et al. (2020). Single-cell transcriptional diversity is a hallmark of developmental potential. Science 367, 405–411. 10.1126/science.aax0249.

14. He, M.X., Cuoco, M.S., Crowdis, J., Bosma-Moody, A., Zhang, Z., Bi, K., Kanodia, A., Su, M.J., Ku, S.Y., Garcia, M.M., et al. (2021). Transcriptional mediators of treatment resistance in lethal prostate cancer. Nat Med 27, 426–433. 10.1038/s41591-021-01244-6.

15. Aspuria, P.P., Lunt, S.Y., Varemo, L., Vergnes, L., Gozo, M., Beach, J.A., Salumbides, B., Reue, K., Wiedemeyer, W.R., Nielsen, J., et al. (2014). Succinate dehydrogenase inhibition leads to epithelial-mesenchymal transition and reprogrammed carbon metabolism. Cancer Metab 2, 21. 10.1186/2049-3002-2-21.

16. Castle, C.D., Cassimere, E.K., Lee, J., and Denicourt, C. (2010). Las1L is a nucleolar protein required for cell proliferation and ribosome biogenesis. Mol Cell Biol 30, 4404–4414. 10.1128/MCB.00358-10.

17. Cheng, L., He, Q., Liu, B., Chen, L., Lv, F., Li, X., Li, Y., Liu, C., Song, Y., and Xing, Y. (2023). SGK2 promotes prostate cancer metastasis by inhibiting ferroptosis via upregulating GPX4. Cell Death Dis 14, 74. 10.1038/s41419-023-05614-5.

18. Honda, M., Nakashima, K., and Katada, S. (2017). PRMT1 regulates astrocytic differentiation of embryonic neural stem/precursor cells. J Neurochem 142, 901–907. 10.1111/jnc.14123.

19. Zhao, Y., Liu, X., He, Z., Niu, X., Shi, W., Ding, J.M., Zhang, L., Yuan, T., Li, A., Yang, W., and Lu, L. (2016). Essential role of proteasomes in maintaining self-renewal in neural progenitor cells. Sci Rep 6, 19752. 10.1038/srep19752.

20. Abida, W., Cyrta, J., Heller, G., Prandi, D., Armenia, J., Coleman, I., Cieslik, M., Benelli, M., Robinson, D., Van Allen, E.M., et al. (2019). Genomic correlates of clinical outcome in advanced prostate cancer. Proc Natl Acad Sci U S A 116, 11428–11436. 10.1073/pnas.1902651116.

21. Grasso, C.S., Wu, Y.M., Robinson, D.R., Cao, X., Dhanasekaran, S.M., Khan, A.P., Quist, M.J., Jing, X., Lonigro, R.J., Brenner, J.C., et al. (2012). The mutational landscape of lethal castration-resistant prostate cancer. Nature 487, 239–243. 10.1038/nature11125.

22. Hubbard, G.K., Mutton, L.N., Khalili, M., McMullin, R.P., Hicks, J.L., Bianchi-Frias, D., Horn, L.A., Kulac, I., Moubarek, M.S., Nelson, P.S., et al. (2016). Combined MYC Activation and Pten Loss Are Sufficient to Create Genomic Instability and Lethal Metastatic Prostate Cancer. Cancer Res 76, 283–292. 10.1158/0008-5472.CAN-14-3280.

23. Kwan, E.M., Dai, C., Fettke, H., Hauser, C., Docanto, M.M., Bukczynska, P., Ng, N., Foroughi, S., Graham, L.K., Mahon, K., et al. (2021). Plasma Cell-Free DNA Profiling of PTEN-PI3K-AKT Pathway Aberrations in Metastatic Castration-Resistant Prostate Cancer. JCO Precis Oncol 5. 10.1200/PO.20.00424.

24. Lin, H.K., Hu, Y.C., Lee, D.K., and Chang, C. (2004). Regulation of androgen receptor signaling by PTEN (phosphatase and tensin homolog deleted on chromosome 10) tumor suppressor through distinct mechanisms in prostate cancer cells. Mol Endocrinol 18, 2409–2423. 10.1210/me.2004-0117.

25. Davies, A.H., Beltran, H., and Zoubeidi, A. (2018). Cellular plasticity and the neuroendocrine phenotype in prostate cancer. Nat Rev Urol 15, 271–286. 10.1038/nrurol.2018.22.

26. Pattabiraman, D.R., Bierie, B., Kober, K.I., Thiru, P., Krall, J.A., Zill, C., Reinhardt, F., Tam, W.L., and Weinberg, R.A. (2016). Activation of PKA leads to mesenchymal-to-epithelial transition and loss of tumor-initiating ability. Science 351, aad3680. 10.1126/science.aad3680.

27. Storrs, E.P., Chati, P., Usmani, A., Sloan, I., Krasnick, B.A., Babbra, R., Harris, P.K., Sachs, C.M., Qaium, F., Chatterjee, D., et al. (2023). High-dimensional deconstruction of pancreatic cancer identifies tumor microenvironmental and developmental stemness features that predict survival. NPJ Precis Oncol 7, 105. 10.1038/s41698-023-00455-z.

28. Takahashi, K., and Yamanaka, S. (2006). Induction of pluripotent stem cells from mouse embryonic and adult fibroblast cultures by defined factors. Cell 126, 663–676. 10.1016/j.cell.2006.07.024.

29. Chaudhuri, A.A., Chabon, J.J., Lovejoy, A.F., Newman, A.M., Stehr, H., Azad, T.D., Khodadoust, M.S., Esfahani, M.S., Liu, C.L., Zhou, L., et al. (2017). Early Detection of Molecular Residual Disease in Localized Lung Cancer by Circulating Tumor DNA Profiling. Cancer Discov 7, 1394–1403. 10.1158/2159-8290.CD-17-0716.

30. Newman, A.M., Bratman, S.V., To, J., Wynne, J.F., Eclov, N.C., Modlin, L.A., Liu, C.L., Neal, J.W., Wakelee, H.A., Merritt, R.E., et al. (2014). An ultrasensitive method for quantitating circulating tumor DNA with broad patient coverage. Nat Med 20, 548–554. 10.1038/nm.3519.

31. Newman, A.M., Lovejoy, A.F., Klass, D.M., Kurtz, D.M., Chabon, J.J., Scherer, F., Stehr, H., Liu, C.L., Bratman, S.V., Say, C., et al. (2016). Integrated digital error suppression for improved detection of circulating tumor DNA. Nat Biotechnol 34, 547–555. 10.1038/nbt.3520.

32. Webster, J., Dang, H.X., Chauhan, P.S., Feng, W., Shiang, A., Harris, P.K., Pachynski, R.K., Chaudhuri, A.A., and Maher, C.A. (2023). PACT: a pipeline for analysis of circulating tumor DNA. Bioinformatics 39. 10.1093/bioinformatics/btad489.

33. Rausch, T., Zichner, T., Schlattl, A., Stutz, A.M., Benes, V., and Korbel, J.O. (2012). DELLY: structural variant discovery by integrated paired-end and split-read analysis. Bioinformatics 28, i333–i339. 10.1093/bioinformatics/bts378.

34. Layer, R.M., Chiang, C., Quinlan, A.R., and Hall, I.M. (2014). LUMPY: a probabilistic framework for structural variant discovery. Genome Biol 15, R84. 10.1186/gb-2014-15-6-r84.

35. Chen, X., Schulz-Trieglaff, O., Shaw, R., Barnes, B., Schlesinger, F., Kallberg, M., Cox, A.J., Kruglyak, S., and Saunders, C.T. (2016). Manta: rapid detection of structural variants and indels for germline and cancer sequencing applications. Bioinformatics 32, 1220–1222. 10.1093/bioinformatics/btv710.

36. Cingolani, P., Platts, A., Wang le, L., Coon, M., Nguyen, T., Wang, L., Land, S.J., Lu, X., and Ruden, D.M. (2012). A program for annotating and predicting the effects of single nucleotide polymorphisms, SnpEff: SNPs in the genome of Drosophila melanogaster strain w1118; iso-2; iso-3. Fly (Austin) 6, 80–92. 10.4161/fly.19695.

37. Talevich, E., Shain, A.H., Botton, T., and Bastian, B.C. (2016). CNVkit: Genome-Wide Copy Number Detection and Visualization from Targeted DNA Sequencing. PLoS Comput Biol 12, e1004873. 10.1371/journal.pcbi.1004873.

38. Cibulskis, K., Lawrence, M.S., Carter, S.L., Sivachenko, A., Jaffe, D., Sougnez, C., Gabriel, S., Meyerson, M., Lander, E.S., and Getz, G. (2013). Sensitive detection of somatic point mutations in impure and heterogeneous cancer samples. Nat Biotechnol 31, 213–219. 10.1038/nbt.2514.

39. Kim, S., Scheffler, K., Halpern, A.L., Bekritsky, M.A., Noh, E., Kallberg, M., Chen, X., Kim, Y., Beyter, D., Krusche, P., and Saunders, C.T. (2018). Strelka2: fast and accurate calling of germline and somatic variants. Nat Methods 15, 591–594. 10.1038/s41592-018-0051-x.

40. Koboldt, D.C., Chen, K., Wylie, T., Larson, D.E., McLellan, M.D., Mardis, E.R., Weinstock, G.M., Wilson, R.K., and Ding, L. (2009). VarScan: variant detection in massively parallel sequencing of individual and pooled samples. Bioinformatics 25, 2283–2285. 10.1093/bioinformatics/btp373.

41. Ye, K., Schulz, M.H., Long, Q., Apweiler, R., and Ning, Z. (2009). Pindel: a pattern growth approach to detect break points of large deletions and medium sized insertions from paired-end short reads. Bioinformatics 25, 2865–2871. 10.1093/bioinformatics/btp394.

42. McKenna, A., Hanna, M., Banks, E., Sivachenko, A., Cibulskis, K., Kernytsky, A., Garimella, K., Altshuler, D., Gabriel, S., Daly, M., and DePristo, M.A. (2010). The Genome Analysis Toolkit: a MapReduce framework for analyzing next-generation DNA sequencing data. Genome Res 20, 1297–1303. 10.1101/gr.107524.110.

43. Ainscough, B.J., Griffith, M., Coffman, A.C., Wagner, A.H., Kunisaki, J., Choudhary, M.N., McMichael, J.F., Fulton, R.S., Wilson, R.K., Griffith, O.L., and Mardis, E.R. (2016). DoCM: a database of curated mutations in cancer. Nat Methods 13, 806–807. 10.1038/nmeth.4000.

44. Tan, A., Abecasis, G.R., and Kang, H.M. (2015). Unified representation of genetic variants. Bioinformatics 31, 2202–2204. 10.1093/bioinformatics/btv112.

45. Khanna, A., Larson, D.E., Srivatsan, S.N., Mosior, M., Abbott, T.E., Kiwala, S., Ley, T.J., Duncavage, E.J., Walter, M.J., Walker, J.R., et al. (2021). Bam-readcount -- rapid generation of basepair-resolution sequence metrics. ArXiv.

46. McLaren, W., Gil, L., Hunt, S.E., Riat, H.S., Ritchie, G.R., Thormann, A., Flicek, P., and Cunningham, F. (2016). The Ensembl Variant Effect Predictor. Genome Biol 17, 122. 10.1186/s13059-016-0974-4.

47. Dodt, M., Roehr, J.T., Ahmed, R., and Dieterich, C. (2012). FLEXBAR-Flexible Barcode and Adapter Processing for Next-Generation Sequencing Platforms. Biology (Basel) 1, 895–905. 10.3390/biology1030895.

48. Li, H., Handsaker, B., Wysoker, A., Fennell, T., Ruan, J., Homer, N., Marth, G., Abecasis, G., Durbin, R., and Genome Project Data Processing, S. (2009). The Sequence Alignment/Map format and SAMtools. Bioinformatics 25, 2078–2079. 10.1093/bioinformatics/btp352.

49. Bie, F., Wang, Z., Li, Y., Guo, W., Hong, Y., Han, T., Lv, F., Yang, S., Li, S., Li, X., et al. (2023). Multimodal analysis of cell-free DNA whole-methylome sequencing for cancer detection and localization. Nat Commun 14, 6042. 10.1038/s41467-023-41774-w.

50. Erger, F., Norling, D., Borchert, D., Leenen, E., Habbig, S., Wiesener, M.S., Bartram, M.P., Wenzel, A., Becker, C., Toliat, M.R., et al. (2020). cfNOMe - A single assay for comprehensive epigenetic analyses of cell-free DNA. Genome Med 12, 54. 10.1186/s13073-020-00750-5.

51. Doebley, A.L., Ko, M., Liao, H., Cruikshank, A.E., Santos, K., Kikawa, C., Hiatt, J.B., Patton, R.D., De Sarkar, N., Collier, K.A., et al. (2022). A framework for clinical cancer subtyping from nucleosome profiling of cell-free DNA. Nat Commun 13, 7475. 10.1038/s41467-022-35076-w.

52. Goldman, M.J., Craft, B., Hastie, M., Repecka, K., McDade, F., Kamath, A., Banerjee, A., Luo, Y., Rogers, D., Brooks, A.N., et al. (2020). Visualizing and interpreting cancer genomics data via the Xena platform. Nat Biotechnol 38, 675–678. 10.1038/s41587-020-0546-8.

53. Chen, J., Bardes, E.E., Aronow, B.J., and Jegga, A.G. (2009). ToppGene Suite for gene list enrichment analysis and candidate gene prioritization. Nucleic Acids Res 37, W305–311. 10.1093/nar/gkp427.

